# Solid stress estimations via intraoperative 3D navigation in patients with brain tumors

**DOI:** 10.1101/2024.11.28.24318104

**Authors:** Hadi T. Nia, Meenal Datta, Ashwin S. Kumar, Saeed Siri, Gino B. Ferraro, Sampurna Chatterjee, Jeffrey M. McHugh, Patrick R. Ng, Timothy R. West, Otto Rapalino, Bryan D. Choi, Brian V. Nahed, Lance L. Munn, Rakesh K. Jain

## Abstract

**Background:** Physical forces exerted by expanding brain tumors - specifically the compressive stresses propagated through solid tissue structures - reduces brain perfusion and neurological function, but heretofore has not been directly measured in patients *in vivo*. Solid stress levels estimated from tumor growth patterns are negatively correlated with neurological performance in patients. We hypothesize that measurements of solid stress can be used to inform clinical management of brain tumors.

**Methods:** We developed an intraoperative technique to quantitatively estimate solid stress and brain replacement by the tumor. In 30 patients we made topographic measurements of brain deformation through the craniotomy site with a neuronavigation system during surgical workflows immediately preceding tumor resection (< 5 minutes in the OR). Utilizing these measurements in conjunction with finite element modeling, we calculated solid stress within the tumor and the brain, and estimated the amount of brain tissue replaced, i.e., lost, by the tumor growth.

**Results:** Mean solid stresses were in the range of 10 to 600 Pa, and the amount of tissue replacement was up to 10% of the brain. Brain tissue loss in patients delineated glioblastoma from brain metastatic tumors, and in mice solid stress was a sensitive biomarker of chemotherapy response.

**Conclusions:** We present here a quantitative approach to intraoperatively measure solid stress in patients that can be readily adopted into standard clinical workflows. Brain tissue loss due to tumor growth is a novel mechanical-based biomarker that, in addition to solid stress, may inform personalized management in future clinical studies in brain cancer.

**Key Points:** Intraoperative and computational technique quantified solid stress and tissue loss in 30 patients Solid stress and tissue loss distinguished tumor types, showing potential as clinical biomarkers

**Importance of the Study:** This study addresses a critical gap, as solid stress has been implicated in tumor progression and treatment resistance but not directly measured in patients with brain cancers before. Here, we present a novel intraoperative technique to quantitatively measure solid stress and brain tissue replacement in brain tumor patients. By combining intraoperative neuro-navigation with finite element modeling, we estimate solid stress and quantify the loss of brain tissue replaced by tumor growth. Importantly, higher tissue replacement was associated with glioblastoma compared to metastatic tumors. In mice, solid stress is a sensitive biomarker of treatment response. These findings establish solid stress and tissue replacement as potential physical biomarkers to inform personalized management of brain tumors. Quantifying these mechanical forces during surgery could help predict patient outcomes and guide clinical decision-making.

## Introduction

Distinct from fluid pressure, solid stress is a mechanical force generated by tumor expansion and transmitted by the solid and elastic components of the tissue (cells and extracellular matrix^1^). This physical hallmark of tumors has been shown to promote tumor invasion, hypoxia, immunosuppression, and treatment resistance ^1-5^. Solid stress is also exerted externally on the surrounding normal tissues ^4,6-9^, which is especially relevant in brain tumors. The rigid skull confines and compounds tumor growth-induced forces, increasing intracranial pressure and the compression of blood vessels and neurons in the surrounding normal brain ^7^. Indeed, in a recent study, we found that retrospective estimates of solid stress – inferred from brain tumor growth patterns – are associated with neurological dysfunction in brain tumor patients ^7^. Interestingly, this effect appears to be independent of fluid accumulation, as we and others have reported that edema does not always correlate with neurological function (e.g., by Karnofsky performance score [KPS])^7,10^.

To further investigate the clinical implications of solid stress in brain tumors, we recently developed methods to measure solid stress in tumors *ex vivo* (in mice and humans) or *in vivo* in mice^*11*^ based on the principle that tissue deformation resulting from the release of elastic energy can be used, in combination with tissue material properties, to estimate the magnitude of solid stress in and around tumors ^4,6,7,12^. However, in vivo measurements of solid stress in patients have not been made to date.

Clinicians have historically noted both fluid (i.e. “edema”) and solid (i.e. “mass effect”) mechanical forces exerted by brain tumors via anecdotal evidence, and frequently evaluate these criteria during diagnosis ^13^. Notably, extracranial brain swelling has been observed during the opening of the dura after craniotomy in approximately 25% of cases ^14^. Based on known mechanopathological phenomena in tumors, we propose that the major contributor to this tissue deformation is solid stress^1^. While peritumoral edema - caused by blood-brain-barrier (BBB) breakdown and elevated interstitial fluid pressure (IFP) - has been measured and investigated as a prognostic/predictive biomarker in brain tumor patients^1, 15^, the clinical implications of solid stress are less understood.

To address this gap in knowledge, we have developed a novel method to measure solid stress intraoperatively in brain tumor patients immediately after craniotomy, prior to opening the dura and prior to tumor resection. By measuring and mapping the extension of brain tissue beyond the dural opening of the skull, we can mathematically estimate solid stress and brain tissue loss on a personalized basis from first principles of solid mechanics. Our results from 30 patients has revealed that volumetrically up to 10% of their brain tissue was replaced by tumor. Interestingly, the brain tissue replaced by tumor is inversely proportional to solid stress, and is significantly increased in glioblastoma over brain metastasis patients. To validate our protocol, we tested it in a simulated craniotomy in mouse models of pediatric (ependymoma), adult (glioblastoma), and secondary (metastatic breast cancer) models of brain tumors. Solid stress measured via craniotomy-induced extra cranial brain herniation in mice is found to be a sensitive biomarker of chemotherapy response in mice. These findings suggest that solid stress may serve as a potential distinguishing biomarker between primary and metastatic brain tumors.

## Materials and Methods

### Ethics Statement

All studies were conducted after informed consent was obtained from each patient prior to their involvement/procedure. We obtained Institutional Review Board approval for all procedures from MGB. All mice were obtained from the Edwin L. Steele Laboratories, Massachusetts General Hospital. Animal protocols were approved by and performed in accordance with the Institutional Animal Care and Use Committees (MGH/HMS) and the Association for Assessment and Accreditation of Laboratory Animal Care International.

### Patient protocols

Prior to the surgery, the patient’s MRI imaging was uploaded to the Brainlab neuronavigation system (BRAINLAB AG, Munich, Germany). The patient’s head was fixed in the Mayfield head holder to restrict any movement, and then a reference array was registered by taking points along the patient’s scalp and face which allowed for 3D registration of the Brainlab neuronavigation system in real-time. The surgery was performed in standard fashion, and the neuronavigation registration was verified for accuracy throughout the operation. After the craniotomy was performed, the neuronavigation was confirmed to identify the location (depth) of the dura matched the pre-operative imaging. At this point, the dura was incised, exposing the underlying brain. As per standard protocol, patients did not receive diuretics.

### Cell lines and animal models

U87 and MGG8 GBM cells were cultured in DMEM media (Sigma-Aldrich) with 10% fetal bovine serum (FBS; Thermo Fischer) and implanted orthotopically in the brains of male and female nude mice (8-12 weeks) as previously described ^6,16^. BT474 breast cancer cells were cultured in RPMI 1640 media (Sigma Aldrich) supplemented with 10% FBS, and implanted orthotopically in the frontal cortex of female nude mice (8-12 weeks) bearing subcutaneous estradiol pellets as previously described ^17^. The EphB2-driven ependymoma cell line Ephb266 was the gift of Dr. Richard Gilbertson. Ephb266 cells were cultured in Neurobasal medium (Invitrogen) containing 2 mM L-glutamine, N2 supplement (Invitrogen), B27 supplement (Invitrogen), 20 ng/ml hrEGF (Invitrogen), 20 ng/ml hrbFGF (Invitrogen) and 50 μg/ml BSA in 5% CO2. Ephb266 cells were injected stereotactically in the cerebella of juvenile FVB male and female mice (aged 6-8 weeks).

### Murine imaging and treatment

#### Microultrasound

Tumor size was visualized by 3-D high-resolution micro-ultrasound (Vevo 2100 system; FUJIFILM VisualSonics) in anesthetized mice through the transparent cranial windows. Ultrasound was also used to measure tumor/brain tissue deformation as a readout of solid stress, following previous methods ^6.^.

#### Treatment

Mice were treated with paclitaxel (10 mg/kg i.p. once weekly), the anti-VEGFR2 antibody DC101 (40 mg/kg i.p. twice weekly), and the anti-Her1/Her2 tyrosine kinase inhibitor Lapatinib (100 mg/kg daily by oral gavage for two weeks as previously described ^18^– as this combination has been shown to have moderate control of BT474 tumors – prior to cranial window removal and brain relaxation.

### Measuring brain deformation in mice

#### Simulated craniotomy

Cranial or cerebellar windows were implanted as previously described ^16,19^. Mice were allowed to recover for 10-14 days after cranial window surgery prior to tumor implantation. Once tumors reached the desired size and/or after treatment, the cranial window was removed to release the stored elastic energy, and the mice were sacrificed to dissipate any fluid pressure. After 15 minutes to allow the tissue to relax and deform, the extent of brain deformation was measured via ultrasound.

#### Post-processing

The 3-D images were exported from the VisualSonics software to MATLAB (The MathWorks) for post-processing of the surface data, followed by export to SolidWorks (Waltham, MA) for 3-D construction of the surfaces into a solid object as previously described ^6^. These data were then exported for the finite element analysis in COMSOL Multiphysics (Burlington, MA) described in detail below.

### Measuring deformation in patients

The Brainlab (Munich) neuronavigational system including a coordinate probe and visualization software was used to measure brain deformation in patients in the operating room during tumor resection surgery immediately following craniotomy. Up to 16 data points are measured on the surface of the dura post craniotomy, to map out the craniotomy boundaries (points 1 to 10), the topology of the deformation (points 11 to 15) as well as the maximum deformation (point 16). These points are registered to the patient’s pre-operative MRI, which are segmented to show the brain and tumor regions. MRI segmentations are transferred to complex geometries implemented in SolidWorks (Dassault Systèmes, Vélizy-Villacoublay, France), which are then converted to 3-D solid stress maps via finite element modeling in COMSOL. Using mathematical modeling, the initial diameter of the tumor that generates the measured deformation level is estimated. When the entire brain tissue is replaced (as opposed to pushed) by the tumor, the craniotomy results in zero deformation; this is equivalent to having the initial tumor diameter in the mathematical modeling equal to the final measured diameter measured in MRI. In another extreme case when the tumor growth results in zero brain replacement, and the brain is instead displaced by the tumor, and we observe the maximum deformation. Representative patient simulation results illustrating the von Mises stress distribution in the tumor and the surrounding brain after and before craniotomy.

#### Geometry

Brain and tumor were simplified as sphere geometries to lower the computational costs and complexity. The radii are calculated based on the volume of the brain and tumor, extracted from the 3D image reconstruction that was previously done in MATLAB and SolidWorks (Dassault Systèmes, Vélizy-Villacoublay, France). In addition to radii, the craniotomy window size and the tumor depth are extracted for each patient and are fed to the FEA package for the parametric study simulations.

#### Materials

Brain and tumor are considered as linear elastic materials. The density and Poisson’s ratio for the brain and the tumor are considered 1,045 kg/m3 and 0.45, respectively. The Young’s modulus for the brain and tumor is subject to the parametric study and varies for each particular patient specific simulations.

#### Finite Element Analysis (FEA) Simulations

The simulations are performed based on the Thermal expansion as a proxy for tumor growth and development. The solid mechanics and heat transfer simulations are coupled in COMSOL Multiphysics to conduct the parametric studies and obtain deformation/displacement values after craniotomy. A fine physics-controlled mesh was used to simulate brain tissue deformation for each patient. We performed the FEA analysis by varying the tumor temperatures which results in different deformation/displacement values ranging from zero (representing maximum brain tissue loss) to maximum deformation (representing zero brain tissue loss).

#### Results

The von Mises stress field before and after the craniotomy and the displacement field after the craniotomy were obtained for every patient-specific simulation.

#### Statistical Analysis

Statistics were performed using Prism (GraphPad Software Inc.). Figure legends depict the statistical test used, and the visualization (e.g., mean with error bars showing standard error of the mean). Differences with p < 0.05 are considered statistically significant.

## Results

### Intraoperative Extracranial Measurements of Brain Deformation Mapping and Finite Element Modeling

We developed an intraoperative workflow to quantitatively and reproducibly map changes in the topography of the brain surface upon opening the dura using 3-D neuronavigation. During brain tumor resection surgery for glioblastoma and brain metastatic patients, once the dura was opened, the neuronavigation platform (BRAINLAB AG, Munich, Germany) was used to acquire 16 templated points at the surface of the brain corresponding to the spatial locations around the perimeter of the craniotomy at the level of the dura (**Fig. 1A-i**). We then registered these data points over the topography of the exposed brain, including the maximum point, to estimate the deformation geometry. The MRI-segmented images from axial, coronal, and sagittal views (**Fig. 1A-ii**) were combined with the Brainlab-gathered coordinates and imported into MATLAB followed by 3D modeling in commercial software SolidWorks (Dassault Systèmes, Vélizy-Villacoublay, France) to obtain the key geometric values needed for subsequent finite element analysis simulations to estimate solid stress and brain replacement by tumor (**Fig. 1A-iii**).

**Figure 1.**
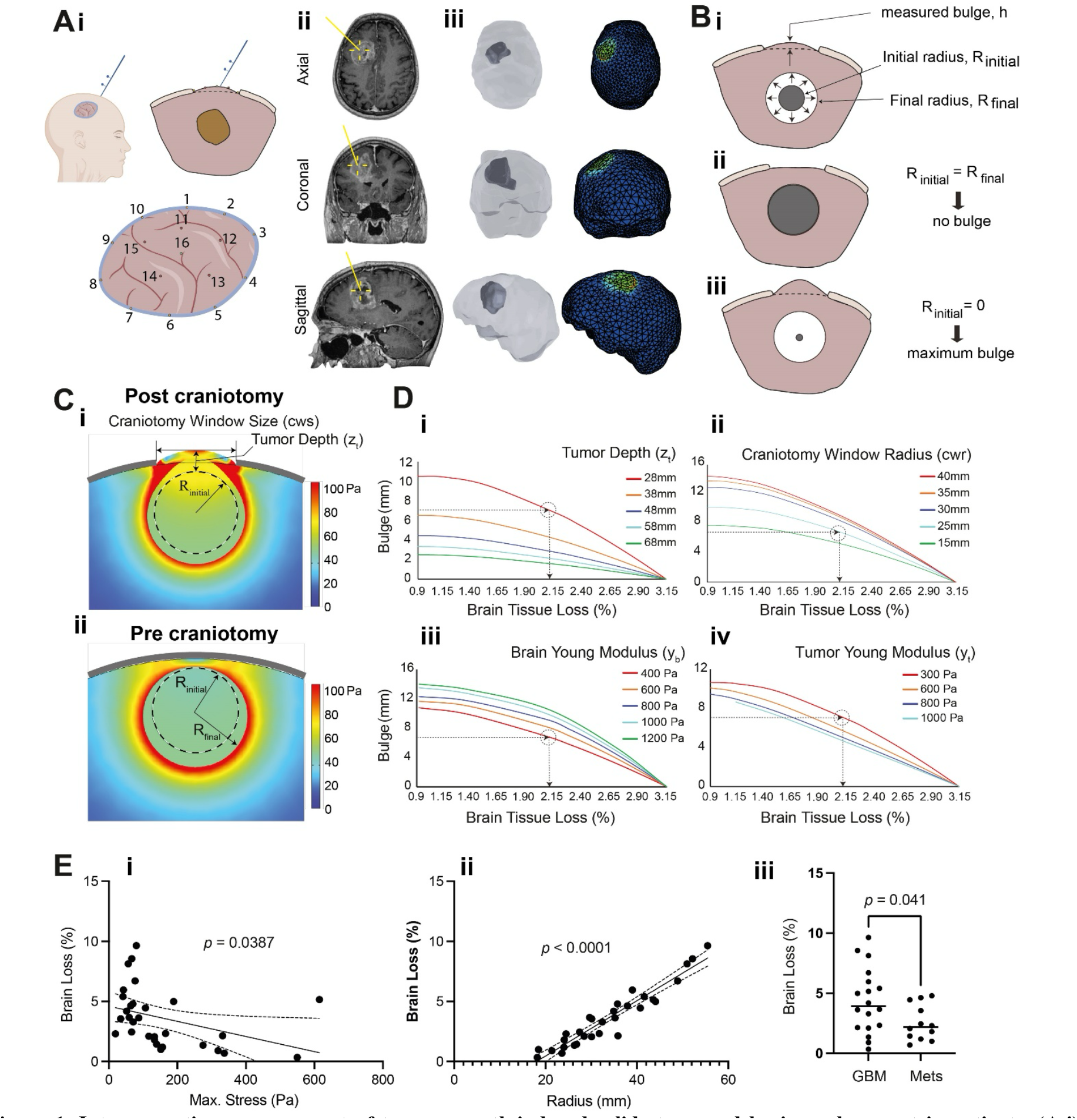
Intra-operative measurement of tumor growth-induced solid stress and brain replacement in patients. (**A-i**) Overview of data collection pipeline to measure solid stress in patients. (**A-ii**) The measurement points are registered to the patient’s pre-operative MRI. (**A-iii**) MRI segmentations are transferred to geometries converted to 3-D solid stress maps via finite element modeling (FEM). (**B-i**) Using FEM, the volume of brain that is replaced by tumor can be estimated. The putative extreme cases of 0% and 100% brain replacement by the tumor are shown in (**B-ii**) and (B-iii), respectively. (**C**) Representative patient simulation results illustrating the stress distribution in the tumor and the surrounding brain (**i**) after and (**ii**) before craniotomy. (**D**) Parametric study showing how the different geometrical and material properties of the tumor-brain affect the post-craniotomy deformation and brain tissue replacement. (**E-i, ii**) Brain replacement is significantly inversely correlated with maximum stress, and is significantly correlated with tumor radius (Pearson’s correlation p-values with regression lines [solid] and 95% confidence interval bands [dashed]). (**E-iii**) Brain replacement can be a potential biomarker to distinguish primary glioblastoma from metastatic tumors, as glioblastoma patients experience significantly higher tissue loss (unpaired Student’s t-test, n=18 glioblastoma [“GBM”], n=12 brain metastasis [“Mets”]; mean ± SEM displayed).

On a per patient basis (with tumor radii ranging from ∼20-45 mm), brain deformation data (**Fig. 1A-i**) combined with tumor and brain dimensions from the pre-operative MRI (**Fig. 1A-ii**) were converted into personalized 3D geometries via computational modeling (in SolidWorks, **Fig. 1A-iii**), from which solid stress was estimated via finite element modeling and first principles of mechanics (in COMSOL, **Fig. 1A-iii**). For the purposes of the finite element analysis, we defined an “initial radius” from which our simulated tumors “grew” to reach the “final radius,” which was defined on a per patient basis from the pre-operative MRI (**Fig. 1B-i**). The initial mathematical radius was estimated so that the deformation that is predicted mathematically matches the deformation measured intraoperatively. This initial mathematical radius is then used to determine the extent of brain tissue loss during tumor growth (displaced vs. replaced). This expansion model uses a mathematical approach that is based on the two extreme scenarios described above. In fact, as shown by the patient data, brain tumors do not recapitulate these two extreme situations, but instead fall within the spectrum of no change in topography (total replacement) vs. maximum change (total displacement; **Fig. 1B-i**).

### Patient-Specific Estimation of Solid Stress and Brain Tissue Replacement

As seen in a typical patient (**Fig. 1C**), the tissue that was originally under compression expands and extends beyond the skull when the solid stress is released via craniotomy. A variety of initial tumor radii were used in the finite element modeling package to simulate various levels of surface deformation until the results converged on the patient-specific topography, tumor depth and size, and craniotomy opening. Here onwards, quantitative estimates of solid stress and brain replacement were made on an individual basis. The solid stress was calculated both intra- and extra-tumorally after solid stress release (**Fig. 1C-i**) and before the craniotomy (**Fig. 1C-ii**). Parametric analyses were also performed to analyze the effects of these variables as well as the material properties (i.e., Young’s Modulus) of the brain and tumor (**Fig. 1D, S3**). These parametric relationships can be used by physicians and scientists to estimate brain tissue replacement based on the input parameter measurements without additional computational steps.

Unlike our previous study, where we inferred that brain tissue may be displaced vs. replaced based on tumor growth patterns (nodular vs. infiltrative, respectively), here we used our measurements of brain deformation after decompression to quantitatively estimate the percent of brain tissue replaced in each patient due to the presence of their tumor. We utilized the following four input parameters from the pre-operative MRI and Brainlab measurements: tumor volume, brain volume, tumor depth from the brain surface, craniotomy (skull hole) dimensions, and brain deformation topology. We found that the 30 patients experienced maximum tissue displacement heights of ∼1-15 mm above the reference plane of the brain, with estimated maximum radial solid stress values of ∼50-600 Pa exerted on the surrounding brain (**Table S1; Figs. S1 and S2**). These patients had 1-10% of their brain tissue replaced by tumor (**Table S1**), which was significantly inversely related to the maximum solid stress values (**Fig. 1E-i**), and significantly positively correlated with tumor size (**Fig. 1E-ii**). Furthermore, patients with glioblastoma (infiltrative, replacing growth phenotype) had higher amounts of brain replacement than patients with brain metastases (nodular, pushing growth phenotype) (**Fig. 1E-iii**). Quantification of individualized brain tissue replacement based on direct tissue deformation measurements in patients has been heretofore unachieved.

### Craniotomy Model in Mice Demonstrates Solid Stress as a Biomarker of Treatment Response

To recapitulate our intraoperative workflow from patients in mouse models, we developed a craniotomy model in mice by removing a previously implanted 7 mm-diameter cranial window (**Fig. 2A**). We used this model to demonstrate the sensitivity of the craniotomy brain deformation as a biomarker of treatment response by treating a Her2+ breast cancer brain metastasis model (BT474). In this model, we treated mice with paclitaxel combined with Her1/2 targeting (Lapatinib) and the anti-angiogenic agent DC101 following our published protocol (**Fig. 2C**) ^18^. We observed a significant reduction in solid stress upon decompression (**Fig. 2Bi)** that was *independent* of a change in average tumor volume (**Fig. 2Bii**). Thus, solid stress is a sensitive biomarker of chemotherapy response in the absence of tumor resection. To demonstrate the versatility of our approach, we also demonstrated the craniotomy model in different models of brain tumors including glioblastoma (nodular U87 vs. infiltrative MGG8) and EphB2+ ependymoma (in the cerebellum, Ephb266 cell line) – via high-resolution ultrasound. The 2-D topographical tissue deformation data was used to estimate 2-D solid stress maps via finite element modeling, and maximum solid stress values co-localize with the tumor site (**Fig. 2D**).

**Figure 2.**
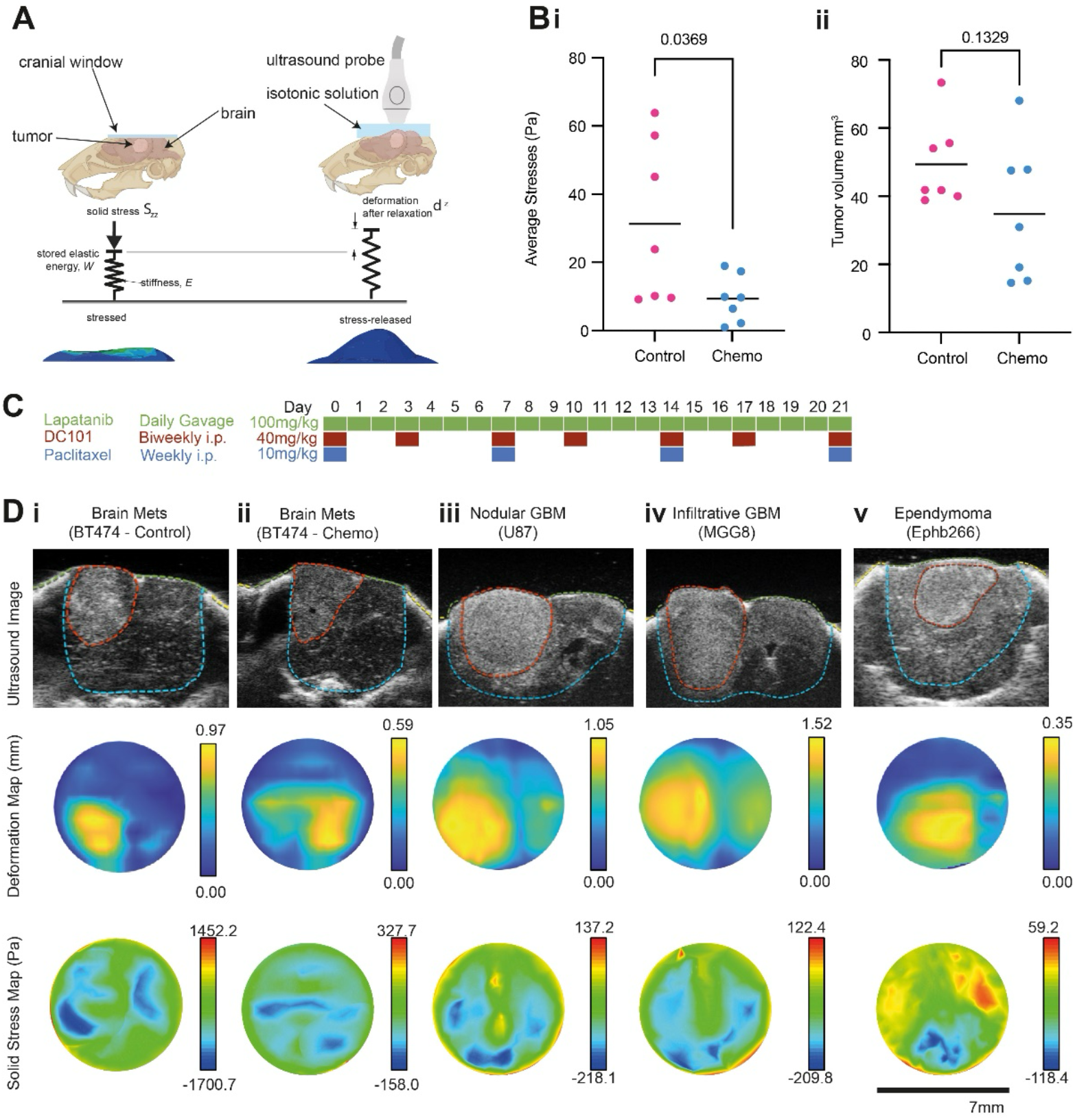
Estimation of solid stress in mouse models of brain tumors, and evaluation of solid stress as a sensitive biomarker of chemotherapy response. (**A**) Solid stress is released and quantified by the removal of the cranial window as a form of simulated craniotomy in mice. After opening the window, the stored elastic energy is released, resulting in brain herniation above the skull in the *z*-direction. (**B**) Solid stress is significantly reduced post-treatment, given that the tumor volume shows only a moderate (non-significant) reduction post-treatment as detailed in (**C**). (**D**) The ultrasound images, 2-D deformation maps, and solid stress maps of Her2+ BT474 breast cancer, nodular U87 glioblastoma (“GBM”), infiltrative MGG8 glioblastoma, and Ephb266 (EphB2+) ependymoma cells.

## Discussion

We provide the first direct measurements of solid stress in brain tumor patients, and a facile framework for clinicians to adopt prospective evaluation of solid stress in their intraoperative procedures. We report the first method to determine the extent of brain tissue replacement (up to 10% in some patients) as a consequence of the tumor disease burden and underlying tumor biology. Both solid stress and tissue loss have the potential to become clinically relevant predictive/prognostic biomarkers for brain tumors.

Solid stress imposed by the tumor on the surrounding parenchyma, clinically deemed as mass effect, can lead to severe neurological consequences. We recently correlated indirect measurements of solid stress – based on brain tumor growth patterns – to Karnofsky Performance Score (KPS) ^7^. Informed by results from murine models that showed solid stress magnitude and effects are dependent on tumor growth patterns, we observed the following in patients: “nodular” tumors displace the surrounding tissue and exert higher forces (worse KPS), while “infiltrative” tumors replace the surrounding tissue and exert lower forces (better KPS). Thus, higher levels of this mechanical biomarker are associated with increased neurological dysfunction.

In our previous study, by relying on radiographic imaging to determine tumor growth patterns, we correlated indirect measurements of solid stress to Karnofsky Performance Score (KPS) ^7^ and made qualitative inferences regarding brain displacement versus brain loss (i.e., tissue replacement). Here, we developed methods to directly measure solid stress during standard intraoperative procedures and to quantitatively estimate a novel biomarker – brain tissue replacement – on an individualized basis. Indeed, by leveraging a surgical navigation system coupled with mathematical modeling, we can objectively and functionally identify growth phenotype (nodular vs. infiltrative) based on brain tissue relaxation and estimate tissue replacement rather than making subjective inferences based on radiographic imaging. Previous simulations of brain deformation and tissue replacement in the presence of a tumor relied on simulated biomechanics and generalized parameters ^20-21^, rather than personalized and directly measured values for each patient as established here.

One of the limitations of the present study is that we neglect the contribution of fluid pressure in our estimations of tissue replacement. In mice, we eliminate this confounding factor by sacrificing the animals prior to making our mechanical measurements; lack of cardiac pumping ceases blood flow, which is the driving force of interstitial fluid pressure (IFP). After a brief period of IFP dissipation to zero, any resulting tissue deformation is attributable to solid stress alone. However, in patients, fluid pressure has been implicated as a cause of brain tissue compression, midline shift, and brain herniation in conditions such as stroke, diabetic ketoacidosis, infection, and acute injury ^22-25^. This pressure is often relieved via decompressive craniectomy ^24,26-27^, which has been mathematically modeled by Kuhl and colleagues ^28-31^. In the case of brain tumors, it is generally accepted that nodular tumors that deform the surrounding tissue (e.g., many brain metastases) have a peritumoral edema that is predominantly vasogenic and fluid dominated ^7,13,32-34^. Infiltrative tumors (e.g., brain-replacing, like many glioblastomas) have a peritumoral edematous area that also contains invading cancer cells^13,32-34^. Recent efforts to distinguish these two cases - namely the presence or lack of cancer cells in the peritumoral edematous area - have been minimally successful with contradictory findings emerging in the literature regardless of imaging method. Baris et al. have defined a “mass-edema index” as the ratio of FLAIR and T2-weighted image area (presumed to be peritumoral edema) to T1-weighted image area (presumed to be tumor volume) ^13^, thus presenting a parameter that circumvents the need to define each imaging modality (T1-vs T2-weighted) as distinctly solid vs fluid, respectively, while He et. al have achieved some success using computationally expensive supervised machine-learning algorithms on a small set of patient MRIs^34^. Advances in imaging technologies that facilitate the differentiation between fluid-dominated peritumoral area vs. cancer cell-containing peritumoral area will allow us to account for the contribution of fluid pressure in subsequent experimental and computational efforts.

In conclusion, our study provides the first *in vivo* measurements of solid stress in brain tumor patients and highlights its potential as a novel biomarker with diagnostic and prognostic value. These findings underscore the importance of considering both fluid and mechanical forces in the clinical management of brain tumors and open new avenues for the development of therapeutic strategies aimed at alleviating the detrimental effects of solid stress on the surrounding normal brain tissue. In the future, quantitative estimates of both solid stress and tissue replacement can be made in prospective trials with brain tumor patients and linked to clinical parameters (e.g., KPS, treatment outcomes, survival) to establish the predictive/prognostic value of these mechanical and personalized biomarkers.

## Data Availability

All data produced in the present study are available upon reasonable request to the authors

## Acknowledgements

We thank the members of the Steele laboratories for their insightful discussions and suggestions. We also thank Carolyn Smith and Sylvie Roberge for their technical assistance.

## Author Contributions

Designing research studies: H.T.N., M.D.; Conducting Animal Experiments: H.T.N., M.D., G.B.F., S.C.; Acquiring Patient Data: J.M.M., P.R.N., T.R.W, O.R., B.D.C., B.V.N.; Analyzing and interpreting data: H.T.N., M.D., A.S.K., S.S., O.R.; Writing Manuscript: H.T.N., M.D.; Reviewing and editing of manuscript: All authors; Supervision of Study: R.K.J., L.M.M., B.V.N.

