## supplementary information for "Solid stress estimations via intraoperative 3D navigation in patients with brain tumors"

**Figures**

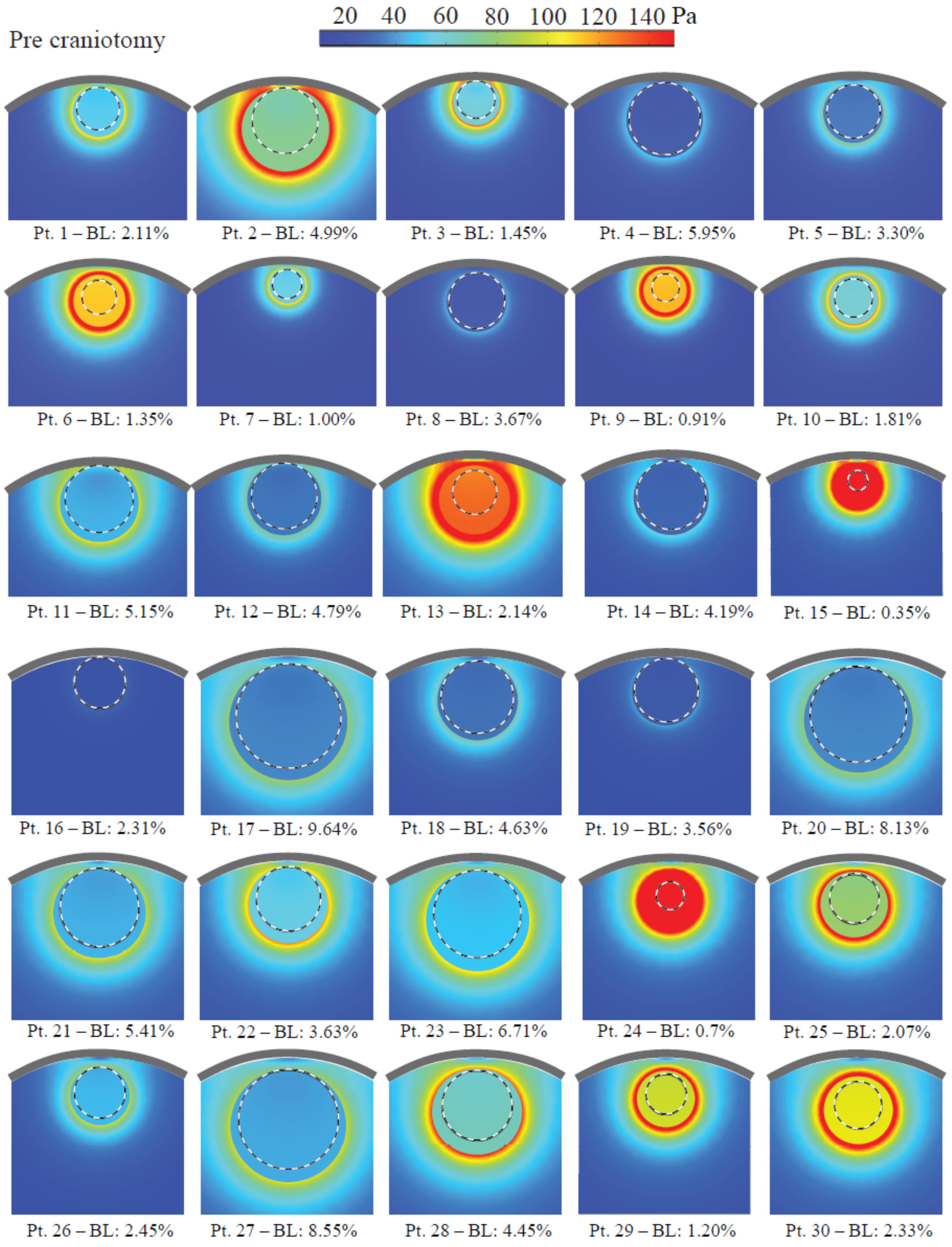


**Figure S1 | The stress map pre-craniotomy in all the individual patients involved in this study.** (BL = brain loss, %.)

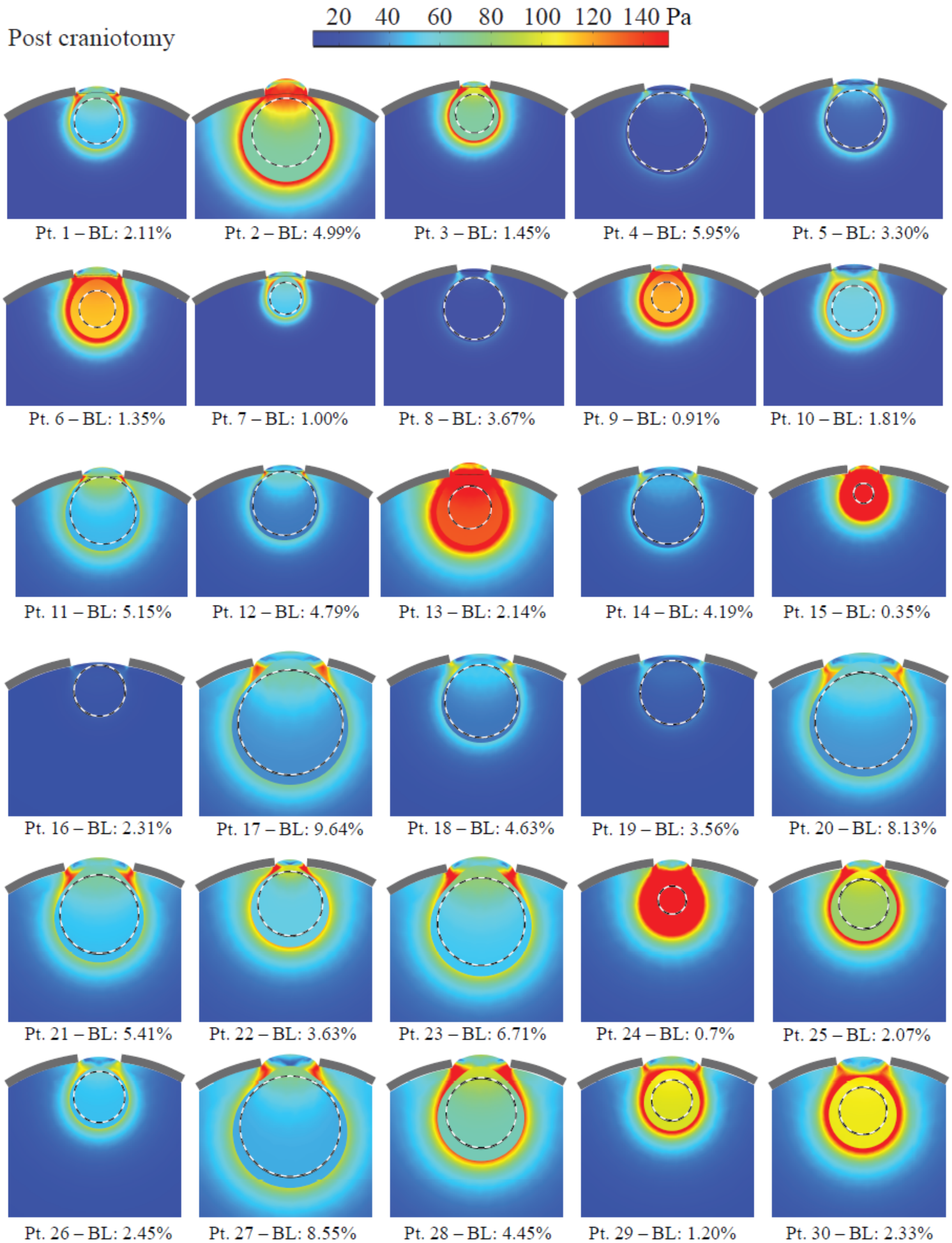


**Figure S2 | The stress map post-craniotomy in all the individual patients involved in this study.** (BL = brain loss, %.)

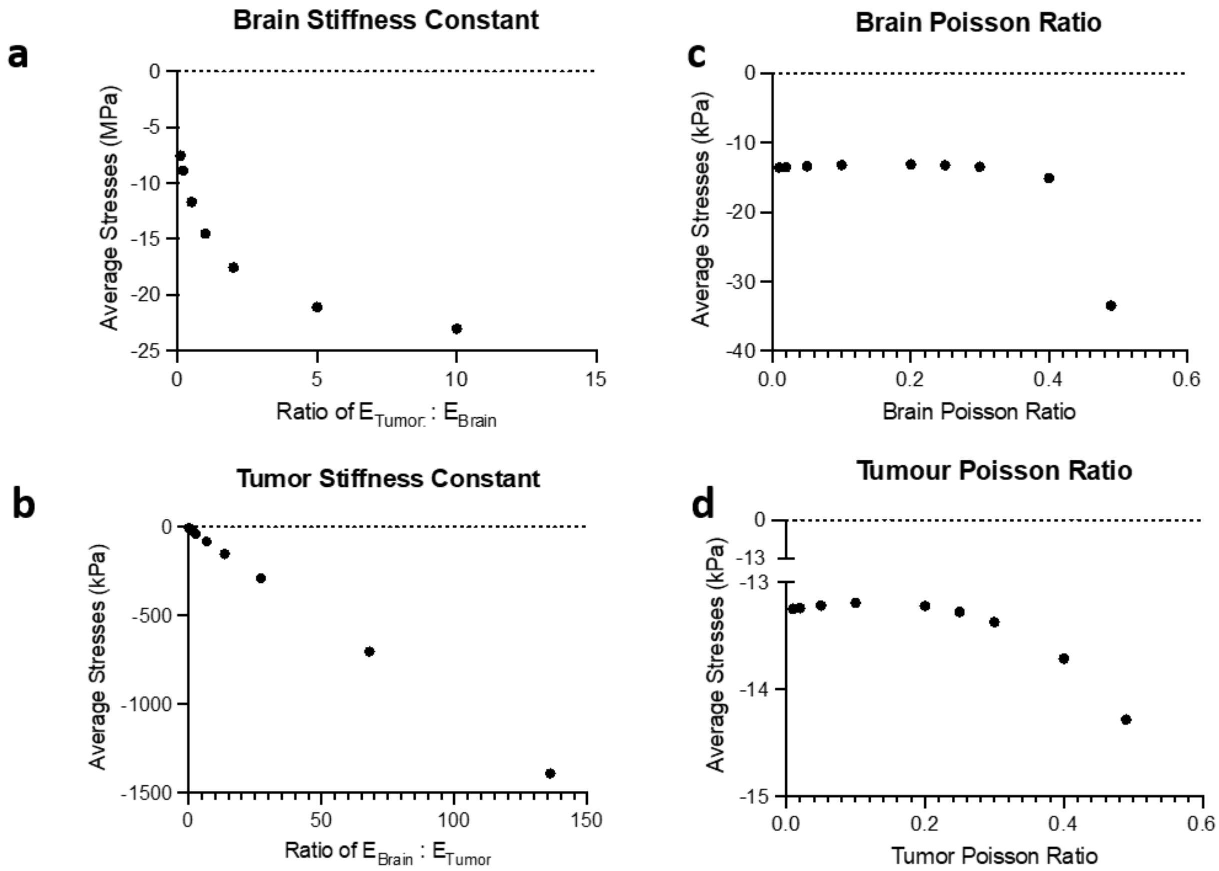


**Figure S3 | Parametric analysis of the effects of tumor and brain material properties on stored solid stresses.**

**Table S1 | Individualized solid stress and brain tissue replacement metrics in patients.**

| patient ID | tumor radius (mm) | max deformation (mm) | max stress (Pa) | tissue replacement (% of brain) |
|---|---|---|---|---|
| pt1 | 28.46 | 6.70 | 116.80 | 2.11 |
| pt2 | 44.09 | 14.47 | 189.14 | 4.99 |
| pt3 | 26.77 | 5.60 | 139.13 | 1.45 |
| pt4 | 39.02 | 3.57 | 42.38 | 5.95 |
| pt5 | 32.43 | 4.56 | 70.70 | 3.30 |
| pt6 | 26.31 | 7.93 | 275.00 | 1.35 |
| pt7 | 18.44 | 4.35 | 151.89 | 1.00 |
| pt8 | 29.64 | 1.82 | 59.39 | 3.67 |
| pt9 | 21.37 | 5.05 | 318.97 | 0.91 |
| pt10 | 24.09 | 3.69 | 133.00 | 1.81 |
| pt11 | 43.59 | 10.08 | 615.00 | 5.15 |
| pt12 | 35.76 | 6.30 | 70.00 | 4.79 |
| pt13 | 35.84 | 14.59 | 332.00 | 2.14 |
| pt14 | 34.90 | 4.86 | 51.90 | 4.19 |
| pt15 | 18.17 | 6.46 | 550.00 | 0.35 |
| pt16 | 24.49 | 1.54 | 19.20 | 2.31 |
| pt17 | 55.48 | 10.55 | 80.30 | 9.64 |
| pt18 | 37.92 | 6.17 | 63.83 | 4.63 |
| pt19 | 30.13 | 3.22 | 34.30 | 3.56 |
| pt20 | 50.98 | 10.68 | 56.52 | 8.13 |
| pt21 | 41.60 | 9.16 | 41.29 | 5.41 |
| pt22 | 35.22 | 6.10 | 87.44 | 3.63 |
| pt23 | 48.92 | 11.32 | 76.55 | 6.71 |
| pt24 | 23.59 | 5.21 | 337.15 | 0.70 |
| pt25 | 30.08 | 6.87 | 133.04 | 2.07 |

| | | | | |
|---|---|---|---|---|
| pt26 | 27.45 | 3.92 | 66.35 | 2.45 |
| pt27 | 52.19 | 8.54 | 66.94 | 8.55 |
| pt28 | 40.76 | 9.88 | 106.94 | 4.45 |
| pt29 | 24.03 | 6.45 | 156.57 | 1.20 |
| pt30 | 31.76 | 6.32 | 165.67 | 2.33 |
